# Acceptance of COVID 19 vaccine among sub-Sahara African (SSA): a comparative study of residents and diaspora dwellers

**DOI:** 10.1101/2022.03.16.22272510

**Authors:** Chundung Asabe Miner, Chikasirimobi G. Timothy, Khathutshelo Percy Mashige, Uchechukwu Levi Osuagwu, Esther Awazzi Envuladu, Onyekachukwu Mary-Anne Amiebenomo, Godwin Ovenseri-Ogbomo, Deborah Donald Charwe, Piwuna Christopher Goson, Bernadine N Ekpenyong, Emmanuel Kwasi Abu, Raymond Langsi, Richard Oloruntoba, Tanko Ishaya, Kingsley Agho

## Abstract

The COVID-19 vaccines are being rolled out across all the Sub-Saharan Africa (SSA) countries, with countries setting targets for achieving full vaccination rates. The aim of this study was to compare the uptake of, resistance and hesitancy to the COVID-19 vaccine between SSA locally residents and in the diaspora. This was a cross-sectional study conducted using a web and paper-based questionnaire to obtain relevant information on COVID-19 vaccine acceptance. The survey items included questions on demography, uptake and planned acceptance or non-acceptance of the COVID-19 vaccines among SSAs. Multinomial logistic regression was used to determine probabilities of outcomes for factors associated with COVID-19 vaccination resistance and hesitancy among SSA respondents residing within and outside Africa. Uptake of COVID-19 vaccines varied among the local (14.2%) and diaspora (25.3%) residents. There was more resistance to COVID-19 vaccine among locals (68.1%) and across the sociodemographic variables of sex [ adjusted Relative Risk (ARR) =0.73, 95% CI; 0.58 – 0.93], primary/less [ARR =0.22, 95% CI; 0.12 – 0.40] and bachelor’s degree [ARR =0.58, 95% CI; 0.43 – 0.77] educational levels, occupation [ARR =0.32, 95% CI; 0.25 - 0.40] and working status [ARR =1.40, 95%CI; 1.06 - 1.84]. COVID-19 vaccine hesitancy was almost similar between locals and diasporas (17.7% and 17.8% respectively) significant only among healthcare workers [ARR =0.46, 95% CI; 0.16 – 1.35] in the diaspora after adjusting for the variables. Similarly, knowledge and perception of COVID-19 vaccine among locals were substantial, but only perception was remarkable to resistance [ARR =0.86, 95% CI; 0.82 – 0.90] and hesitancy [ARR =0.85, 95% CI; 0.80 – 0.90] of the vaccine. Differences exist in the factors that influence COVID-19 vaccine acceptance between local SSA residents and those in the diaspora. Knowledge about COVID-19 vaccines affects the uptake, resistance, and hesitancy to the COVID-19 vaccine. Information campaigns focusing on the efficacy and safety of vaccines could lead to improved acceptance of COVID-19 vaccines.

## Introduction

The coronavirus disease (COVID-19) pandemic that started in December of 2019, initially reported in Wuhan, China, has continued despite preventative measures adopted worldwide under the guidance of the World Health Organization (WHO). Many countries have experienced their second, third and fourth waves in terms of cases and resultant deaths.[1-4] The outbreak of the new Omicron variant in different countries [5-7] is of global concern,[8] as it threatens the return to normalcy and the ongoing COVID-19 vaccination programmes. Non-pharmaceutical interventions to minimise the spread of infections included travel restrictions, lockdowns, physical distancing, regular handwashing and wearing of face masks.[9-10] From the onset of the pandemic, scientists and pharmaceutical companies began the development of COVID-19 vaccines to offer protection against severe disease.[11]

The Pfizer/BioNTech, Moderna, AstraZeneca/Oxford, Johnson &Johnson, Sinopharm/BIBP and India’s Covishield,[12-13] vaccines are licensed for use across the globe. The utilisation of any vaccine can be influenced by system, client and provider factors,[14] but in particular, vaccine acceptance plays a huge role for clients and providers. Generally, the acceptance of any vaccine has been shown to be influenced by demographic factors, knowledge of the disease and the consequences of contracting it, perceptions of susceptibility, potential benefits of a health action and the occurrence of one or more cues to action.[11-17] Similar factors may influence COVID-19 vaccine acceptance.

The COVID-19 vaccines have shown to be efficient and safe,[18] however, their acceptance is a major barrier to the successful rollout plans in different countries including the SSA region. This is further exacerbated by the mistrust in the government demonstrated by residents in this region.[19] The WHO defines vaccine hesitancy as a ‘delay in acceptance or refusal of safe vaccines despite availability of vaccine services’[20] It is also stated to be one of the top ten threats to global health.[21-22] Vaccine hesitancy is used to describe a phenomenon where individuals are unsure of getting vaccinated. Those who object to getting the vaccine are defined as vaccine resistant. [23]

The success of vaccines depends on achieving maximum coverage and thereby attaining herd immunity.[24] Vaccine acceptance is therefore crucial to the efforts currently made by public health experts of ensuring that the communities in every country are fully vaccinated. Studies have shown that there have been disparities in vaccine acceptance for other conditions, and factors such as age, race and ethnicity, social class, country and region of origin were associated with acceptance of vaccines.[25-26] Similar results were reported for COVID-19 vaccines.[27-28]

Persons in the diaspora are “national migrant communities living in interaction among themselves and with their country of origin”.[29] Africans in the diaspora have been referred to by the African Union as “people of African origin living outside of the continent, irrespective of their citizenship and nationality, and who are willing to contribute to the development of the continent and the building of the African Union”.[30] It is generally believed that being in the diaspora provides Africans with greater opportunities to become more enlightened and therefore adopt different approaches to decision making.[30] Furthermore, studies have shown that there is geographical and spatial variation in the uptake of vaccines.[31-32] In SSA, access to COVID-19 vaccines have improved, but the availability of vaccines and uptake remains substantially low compared with the rich European and North-American countries [33], and only 11% of the adult population in Africa are fully vaccinated as at January 2021.[34] Although there are significant differences in the vaccination programmes and their rollout between countries,[35-36] the fact that a previous study found similarities in the attitude and risk perception towards COVID-19 among Africans living locally and those in the diaspora (mostly living in Western countries) during the lockdown, [37] suggests there could be similarities in their acceptance of the COVID-19 vaccination. This study, therefore, sought to investigate the differences in the acceptance of COVID-19 vaccination of Sub-Sahara Africans living on the African continent and those in the diaspora. Although different studies exist that looked at COVID-19 vaccine acceptance, none had compared the same between locals and diaspora dwellers in SSA at the time of this study.

## Methods

### Ethics and consent

Ethical approval to conduct the study was obtained from the Humanities and Social Sciences Research Ethics Committee (approval #: HSSREC 00002504/2021) of the University of KwaZulu-Natal, Durban, South Africa. The study adhered to the tenets of the Declaration of Helsinki involving human participants [38], and anonymous voluntary informed consent was obtained from all participants as part of the preamble accompanying the questionnaire.

Participants were included in this study if they were of African origin, aged 18 years and older, and provided consent. Completion of the questionnaire was only possible after the participants had responded to the consent question, ‘do you voluntarily take part in this study?’ Those who answered ‘No’ to this question were automatically locked out from the survey platform.

### Study setting and population

The study population included adults who were 18 years and older and were of sub-Saharan Africans residing locally (in Africa) and in diaspora (outside of Africa). Respondents from several countries in SSA, mostly from Cameroun, Ghana, Nigeria, South Africa, Tanzania, and those in diaspora mostly living in Australia, United Kingdom, United States, Saudi Arabia, Canada, China, and India took part in this study.

### Sample size determination

The sample size was determined using Cochran’s formulae (n = z^2^pq/d^2^) with the assumption of a proportion of 50% at a confidence level of 95% with an error margin of 2.5%. A 20% non-response rate was assumed, and a minimum sample size of 2401 was obtained.

### Study design

This was a web-based cross-sectional survey carried out between 14^th^ of March and 17^th^ of May 2021. Due to the continued COVID-19 lockdowns in many of the target countries at the time of this study, web-based study was most appropriate even though it may have excluded some participants with no access to internet-based phone/computer services.

### The survey instrument and data collection

Data was collected using a validated self-administered questionnaire adapted from a previous study.[39] The survey tool was tested for the internal validity of the items, and Cronbach’s alpha coefficient score ranged from 0.70 and 0.74, indicating satisfactory consistency.[40] The questionnaire was designed on survey monkey in both English and French, which are spoken languages in 26 and 21 SSA countries, respectively.[41] The questionnaire was disseminated electronically through an e-link on social media networks such as WhatsApp, Facebook and e-mail. There was an accompanying introductory section that included the background and goal of the study, procedure for participation and informed consent guide. Participants were requested on the introductory page not to participate in the survey more than once.

### Confounding variables

The survey instrument showing the various variables collected has been presented in the Supporting information (S1 Table). The independent variables included sociodemographic variables; age, gender, region, marital status, the highest level of education, occupation, employment status, religion, smoking status, previous vaccination for other conditions and pre-existing medical conditions; knowledge of COVID-19 vaccines; perception of risk for contracting COVID-19; and attitude towards vaccination for COVID-19 (S1 Table). The exposure variable was the ‘place of residence’ (local or diaspora).

The COVID-19 vaccine knowledge items had 10 questions on a Likert scale with five levels as indicated in SI Table 1. The scores for nine of the items ranged from 0 (lowest) to 4 (highest) while, for one item, it was coded as 1 for Yes and 0 for No. The overall knowledge towards COVID-19 vaccination score ranged from 0 -37 points, with a higher knowledge score indicating a better knowledge towards COVID-19 vaccination.

**Table 1:** Characteristics (n=2545) of the study participants living in (Local) and outside of Africa (Diaspora).

| Variables | Local<br>2391 (93.9%) | Diaspora<br>154 (6.1) |
| --- | --- | --- |
| **Age group (years)** |  |  |
| 18 – 28 | 898 (38.7) | 23 (14.9) |
| 29 – 38 | 677 (29.1) | 41 (26.6) |
| 39 – 48 | 450 (19.4) | 46 (29.9) |
| 49 + | 297 (12.8) | 44 (28.6) |
| **Sex** |  |  |
| Males | 1,264 (52.9) | 112 (72.7) |
| Females | 1,127 (47.1) | 42 (27.7) |
| **SSA region of origin\*** |  |  |
| West Africa | 1,330 (55.6) | 107 (75.4) |
| East Africa | 116 (4.9) | 6 (4.2) |
| Central Africa | 288 (12.1) | 24 (16.9) |
| Southern Africa | 657 (27.5) | 5 (3.5) |
| **Marital status** |  |  |
| Married | 1,030 (43.1) | 92 (59.7) |
| Not married ‡ | 1,361 (56.9) | 62 (40.3) |
| **Highest level of education** |  |  |
| Postgraduate degree (Masters/PhD) | 668 (27.9) | 82 (53.3) |
| Bachelor's degree | 1,237 (51.7) | 61 (39.6) |
| Secondary/High School | 436 (18.2) | 9 (5.8) |
| Primary or Less | 50 (2.1) | 2 (1.3) |
| **Employment status** |  |  |
| Employed/Self employed | 1,733 (72.5) | 139 (90.3) |
| Unemployed/Retired | 658 (27.5) | 15 (9.7) |
| **Religion** |  |  |
| Christianity | 2,140 (89.5) | 138 (89.6) |
| Others | 251 (10.5) | 16 (10.4) |
| **Occupation** |  |  |
| Non-Healthcare | 1,658 (69.3) | 95 (61.7) |
| Healthcare | 733 (30.7) | 59 (38.3) |
| **Previous vaccination for any condition** |  |  |
| No | 430 (18.0) | 15 (9.7) |
| Yes | 1,961 (82.0) | 139 (90.3) |
| **Smoking Status** |  |  |
| Ex-smoker | 142 (5.9) | 18 (11.7) |
| Current smoker | 168 (7.0) | 8(5.2) |
| Non smoker | 2,081 (87.0) | 128 (83.1) |
| **Risk factors: Any pre-existing condition§** |  |  |
| No | 2,022 (84.6) | 107 (69.5) |
| Yes | 369 (15.4) | 47 (30.5) |

*Data presented as frequencies (percentages)*
*\*SSA; Sub-Saharan Africa, ‡ includes single, divorced, and widowed*
*§includes the presence of any of the following conditions: cancer, diabetes, hypertension, asthma, kidney disease, any heart condition, sickle cell anemia*

The attitude towards the COVID-19 vaccine items included four items with each assigned 2 points for ‘yes’, 1 point for ‘unsure’ and 0 point for ‘No’. The total attitude score ranged from 0 to 8, with a higher score denoting a better attitude towards COVID-19 vaccination.

The risk perception for contracting the disease after vaccination included questions on how the participants rate their risk of becoming infected with the virus and risk of dying from the infection. The responses were structured using a Likert scale with five levels (S1 Table), with scores for each item ranging from 0 (lowest) to 4 (highest). The total perception score ranged from 0 to 8, with a higher score representing a higher perception of contracting the infection following COVID-19 vaccination.

### Main outcome variables

The main outcomes were vaccine uptake, resistance and hesitancy. Uptake was determined by answering ‘yes’ to the question “*Have you been vaccinated against COVID-19?*”. The vaccine resistant group were those that answered ‘no’ to the question ‘*Will you be willing to be vaccinated against COVID-19 if the vaccine becomes available in your country?*’, while those who answered ‘not sure’ were defined as the vaccine ‘hesitant’ group.

### Data analysis

Data were analyzed using STATA/MP version 14 (Stata Corp 2015, College Station, TX, USA. A 95% confidence interval (CI) was set for this survey, and a p-value of <0.05 was considered statistically significant. Descriptive data were summarized and presented in tables and charts using frequencies, percentages, mean and standard deviations as required. Multinomial logistic regression analyses were used to examine the COVID-19 vaccination status on sources of information. As part of the multiple multinomial logistic regression analyses, a staged modelling technique was carried out. Elimination method was conducted using multiple multinomial logistic regression modelling techniques to remove statistically non-significant variables. Demographic factors were first entered into the baseline multiple regression model, followed by health indicators factors and the exposure variables were examined in the final model, which also included knowledge, attitude and risk perception variables, keeping only those variables significant in the previous model. In the final model, we tested and reported any co-linearity. The relative risk with 95% confidence intervals were calculated to assess the adjusted risks of independent variables.

## Results

### Characteristics of the respondents

There was a total of 2545 SSA respondents [2391 locals (93.9%) and 154 in the diaspora (6.1%)]. Table 1 shows the frequency and percentage distribution of respondents according to their socio-demographic variables. The majority of the SSA local residents (67.8%) were younger than 38 years, while those in the diaspora were older. There were more females than males in both groups, and the majority were originally from West Africa (locals 55.6%, diaspora 75.4%). More than half (56.9%) of the locals were not married, and 59.7% from the diaspora were married. Many locals had a bachelors’ degree (56.9%), and most diaspora participants were postgraduate degree holders (53.3%). Most respondents from both groups were employed /self-employed were predominantly non-healthcare workers and were of the Christian faith. More than 80% of locals and above 90% of those in the diaspora had been previously vaccinated for one or two other conditions. More than two-thirds of the respondents indicated that they have never smoked. The proportion of respondents with preexisting conditions was high among locals (84.6%) and diaspora (69.5%).

### Prevalence of uptake, resistance and hesitancy towards COVID-19 vaccine in SSA

Figure 1 presents the prevalence of vaccine uptake, resistance and hesitancy in both locals and those in the diaspora. The prevalence of COVID-19 vaccine uptake respondents was almost twice higher among the diaspora (25.3%) than among the locals (14.2%). Resistance to the COVID-19 vaccine was more common among the locals (68.1%) than those in the diaspora (55.2%). Hesitancy to COVID-19 vaccine was almost the same for both locals and resident in the diaspora(See Figure 1).

**Fig I:**
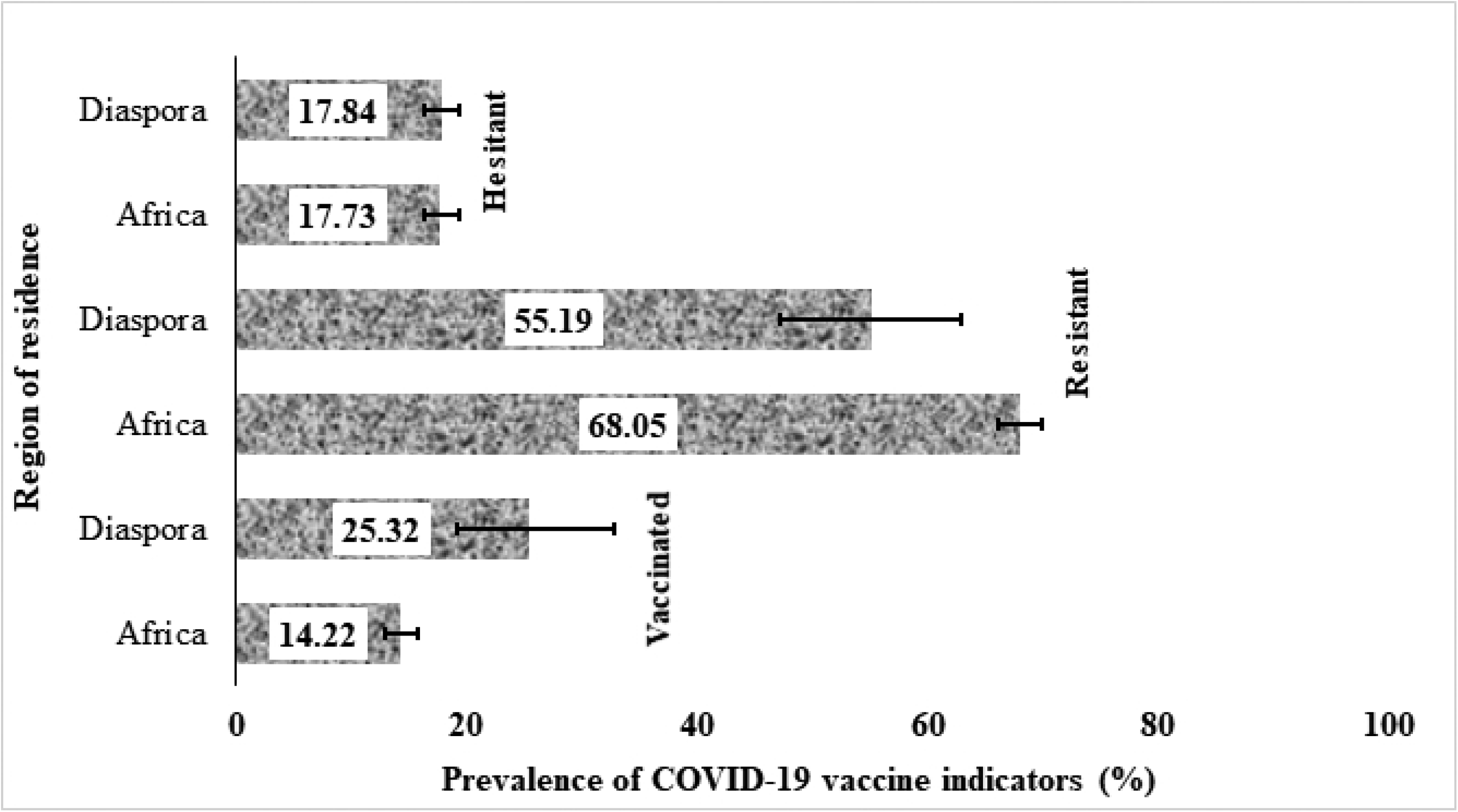
Prevalence and 95% confidence intervals of vaccine uptake, resistance and hesitancy among SSAs living in (within) and outside of Africa (diaspora).

### Distribution of vaccine uptake, resistance and hesitancy among local and diaspora residents

Table 2 shows the variations in the distribution of vaccine uptake, resistance and hesitancy across the demographic variables as well as their mean scores for knowledge, attitude and perception of risk of infection. Those aged between 39 – 48 years had the highest proportion of locals that were resistant to the vaccine (70.0%) while among those in the diaspora, the 18 – 28 years’ age range had the highest proportion (73.9%). More males (70.2%) than females (65.7%) were resistant to taking the vaccine among the locals, whereas there was a preponderance of resistant females in the diaspora group (64.3%).

**Table 2.** Prevalence of vaccine uptake, hesitancy and resistance among SSAs living in (local) and outside Africa (diaspora).

| Variable | LOCAL |  |  | DIASPORA |  |  |
| --- | --- | --- | --- | --- | --- | --- |
|  | Uptake<br>n = 340 | Resistant<br>n = 1627 | Hesitant<br>n = 424 | Uptake<br>n = 39 | Resistant<br>n = 85 | Hesitant<br>n = 30 |
| **Age in years** |  |  |  |  |  |  |
| 18 – 28 | 127 (14.1) | 621 (69.2) | 150 (16.7) | 2 (8.7) | 17 (73.9) | 4 (17.0) |
| 29 – 38 | 101 (14.9) | 435 (64.3) | 141 (20.8) | 3 (7.3) | 25 (61.0) | 13 (31.0) |
| 39 – 48 | 61 (13.6) | 315 (70.0) | 74 (16.4) | 18 (39.1) | 20 (43.5) | 8 (17.0) |
| 49+ | 33 (11.1) | 205 (69.0) | 59 (19.9) | 16 (36.4) | 23 (52.3) | 5 (11.0) |
| **Sex** |  |  |  |  |  |  |
| Males | 159 (12.6) | 887 (70.2) | 218 (17.3) | 32 (28.6) | 58 (51.8) | 22 (19.6) |
| Females | 181 (16.1) | 740 (65.7) | 206 (18.3) | 7 (16.7) | 27 (64.3) | 8 (19.1) |
| **Region** |  |  |  |  |  |  |
| West Africa | 205 (15.4) | 897 (67.4) | 228 (17.1) | 32 (29.9) | 54 (50.5) | 21 (19.6) |
| East Africa | 6 (5.2) | 80 (69.0) | 30 (25.9) | 0 (0.0) | 3 (50.0) | 3 (50.0) |
| Central Africa | 58 (20.1) | 186 (64.6) | 44 (15.3) | 6 (25.0) | 16 (66.7) | 2 (8.3) |
| Southern Africa | 71 (10.8) | 464 (70.6) | 122 (18.6) | 0 (0.0) | 4 (80.0) | 1 (20.0) |
| **Level of education** |  |  |  |  |  |  |
| Master's degree and higher | 68 (10.2) | 472 (70.7) | 128 (19.2) | 22 (26.8) | 42 (51.2) | 18 (22.0) |
| Bachelor's degree | 204 (16.5) | 815 (65.9) | 218 (17.6) | 14 (23.0) | 38 (62.3) | 9 (14.8) |
| Secondary/high school | 49 (11.2) | 311 (71.3) | 76 (17.4) | 3 (33.3) | 4 (44.4) | 2 (22.2) |
| Primary/no school | 19 (38.0) | 29 (58.0) | 2 (4.0) | 0 (0.0) | 1 (50.0) | 1 (50.0) |
| **Occupation** |  |  |  |  |  |  |
| Non-healthcare | 157 (9.0) | 1,188 (71.2) | 313 (18.9) | 17 (17.9) | 61 (64.2) | 17 (17.9) |
| Healthcare | 183 (25.0) | 439 (59.9) | 111 (15.1) | 22 (37.3) | 24 (40.7) | 13 (19.5) |
| **Working status** |  |  |  |  |  |  |
| Employed | 262 (15.1) | 1,149 (66.3) | 322 (18.5) | 38 (27.3) | 76 (54.7) | 25 (18.0) |
| Unemployed | 78 (11.9) | 478 (72.6) | 102 (15.5) | 1 (6.7) | 9 (60.0) | 5 (33.3) |
| **Marital/family status** |  |  |  |  |  |  |
| Married | 126 (12.2) | 719 (69.8) | 185 (18.0) | 29 (31.5) | 47 (51.1) | 16 (17.4) |
| Not married ‡ | 214 (15.7) | 908 (66.7) | 239 (17.6) | 10 (16.1) | 38 (61.3) | 14 (22.6) |
| **Religion** |  |  |  |  |  |  |
| Christians | 304 (14.2) | 1,439 (67.2) | 397 (18.6) | 35 (25.4) | 78 (56.5) | 25 (18.1) |
| Others | 36 (14.3) | 188 (74.9) | 27 (10.8) | 4 (25.0) | 7 (43.8) | 5 (31.3) |
| **Smoking status** |  |  |  |  |  |  |
| Ex-smoker | 21 (14.8) | 98 (69.0) | 23 (16.2) | 4 (22.2) | 13 (72.2) | 1 (5.6) |
| **Current smoker** | 20 (11.9) | 113 (67.3) | 35 (20.8) | 1 (12.5) | 5 (62.5) | 2 (25.0) |
| **Non-smoker** | 299 (14.4) | 1,627 (68.1) | 424 (17.7) | 34 (26.6) | 67 (52.3) | 27 (21.1) |
| **Have you been vaccinated for any condition** |  |  |  |  |  |  |
| **No** | 30 (7.0) | 326 (75.8) | 74 (17.2) | 3 (20.0) | 9 (60.0) | 3 (20.0) |
| **Yes** | 310 (14.2) | 1,301 (66.3) | 350 (17.9) | 36 (25.9) | 76 (54.7) | 27 (19.4) |
| **Any pre-existing conditions<sup>§</sup>** |  |  |  |  |  |  |
| **No** | 280 (13.9) | 1,383 (68.4) | 359 (17.6) | 23 (21.5) | 61 (57.0) | 23 (21.5) |
| **Yes** | 60 (16.3) | 244 (66.1) | 65 (17.6) | 16 (34.0) | 24 (51.1) | 7 (14.9) |
| **Knowledge\*** | 18.7 ± 4.9 | 18.5 ± 6.3 | 19.6 ± 3.6 | 22.7 ± 3.4 | 18.9 ± 6.0 | 20.5 ± 3.8 |
| **Attitude\*** | 1.2 ± 2.2 | 0.9 ± 2.1 | 1.0 ± 2.0 | 0.7 ± 1.9 | 0.7 ± 1.8 | 1.3 ± 2.2 |
| **Perception\*** | 6.7 ± 2.7 | 5.6 ± 3.1 | 5.8 ± 2.3 | 7.2 ± 2.4 | 5.3 ± 3.1 | 5.9 ± 2.9 |

*Data presented in frequencies (percentages). \*Data presented as mean ± standard deviation.*
<sup>‡</sup> includes single, divorced and widowed. <sup>§</sup> includes the presence of any of the following conditions: cancer, diabetes, hypertension, asthma, kidney disease, any heart condition, sickle cell anemia.

COVID-19 vaccine uptake was highest among Central African residents (20.1%) who lived locally but was highest among West Africans (29.9%) in the diaspora. The uptake of COVID-19 vaccine had the highest proportion among those with primary/less education [19 (38%)] while among those in the diaspora, uptake was highest in those having a Master’s degree and higher [22 (26.8%)]. Resistance was substantial in those with Master’s and higher degree respondents (70.7% for locals and 51.2% for those in the diaspora). For both healthcare and non-healthcare workers in both groups, the greatest proportions were resistant to taking the vaccine. The proportion of uptake, hesitancy and resistance towards COVID-19 vaccines varied with the employment status of the respondents, though the unemployed had the highest proportions of vaccine resistance in both groups (72.6% for locals and 60.0% for the diaspora). Christians represented the higher number of those who said they were hesitant to take the vaccine (18.6%) as compared with non-Christians in the diaspora (31.3%). Those who were ex-smokers had the highest proportion of those who were resistant among both the locals (69.0%) and those in the diaspora (72.2%). The uptake of the vaccine was also higher among those with pre-existing conditions in both local and diaspora respondents.

Higher mean scores for attitude and perception were observed among the COVID-19 vaccine uptake respondents for the local residents, while the mean knowledge score was highest for the hesitant group. Among the diasporas, the mean knowledge and perception scores were similarly highest in uptake respondents, but a higher score for attitude was observed in the hesitancy respondents (Table 2).

### Unadjusted analysis of factors associated with COVID-19 vaccine uptake, resistance and hesitancy in SSA

Table 3 shows the unadjusted relative risk of factors associated with resistance and hesitancy towards COVID-19 vaccination among SSA respondents living locally and in the diaspora. Among the local residents, female sex was associated with the COVID-19 vaccine resistance [RR=0.73, 95% CI; 0.58 – 0.93]. East and Southern Africa local residents were significantly associated with COVID-19 vaccine resistance [RR=3.05, 95% CI; 1.31 - 7.08 and RR=1.49, 95% CI; 1.12 - 2.00 respectively] and hesitancy [RR=4.50, 95% CI; 1.83 - 11.02 and RR=1.54, 95% CI; 1.09 - 2.19 respectively]. Having primary or less education was also shown to be significantly associated with COVID-19 vaccine resistance [RR=0.22, 95% CI; 0.12 – 0.41] and hesitancy [RR=0.06, 95% CI; 0.01 – 0.25] among local residents. Unemployment was significantly associated with higher risk of vaccine resistance [RR=1.40, 95% CI; 1.06 – 1.84] among local residents. Being unmarried [RR=0.74, 95% CI; 0.58 – 0.95], and having a history of vaccination for other conditions were associated with lower risk of vaccine resistance [RR=0.39, 95% CI; 0.26 – 0.57] among locals. Also, those with high risk perception scores were significantly less likely to resist [RR=0.88, 95% CI; 0.84 - 0.92] or be hesitant [RR=0.90, 95% CI; 0.85 - 0.94] to the COVID-19 vaccines.

**Table 3.** Relative risk (RR) for factors associated with COVID-19 vaccine uptake, hesitancy and resistance among SSA locals and diasporas. The base reference was COVID-19 vaccine uptake for all variables.

| Variable | LOCAL |  | DIASPORA |  |
| --- | --- | --- | --- | --- |
|  | Resistant<br>RR (95%CI) | Hesitant<br>RR (95%CI) | Resistant<br>RR (95%CI) | Hesitant<br>RR (95%CI) |
| **Age in years** |  |  |  |  |
| 18 – 28 | 1.00 | 1.00 | 1.00 | 1.00 |
| 29 – 38 | 0.88 (0.66 - 1.18) | 1.18 (0.83 - 1.67) | 0.98 (0.15 - 6.50) | 2.17 (0.26 - 17.89) |
| 39 – 48 | 1.06 (0.76 - 1.47) | 1.03 (0.68 - 1.55) | 0.13 (0.03 - 0.65) | 0.22 (0.03 - 1.47) |
| 49+ | 1.27 (0.84 - 1.92) | 1.51 (0.93 - 2.46) | 0.17 (0.03 - 0.84) | 0.16 (0.02 - 1.12) |
| **Sex** |  |  |  |  |
| Males | 1.00 | 1.00 | 1.00 | 1.00 |
| Females | 0.73 (0.58 - 0.93) | 0.83 (0.62 - 1.10) | 2.13 (0.83 - 5.43) | 1.66 (0.53 - 5.25) |
| **Region** |  |  |  |  |
| West Africa | 1.00 | 1.00 | 1.00 | 1.00 |
| East Africa | 3.05 (1.31 - 7.08) | 4.50 (1.83 - 11.02) | - | - |
| Central Africa | 0.73 (0.53 - 1.02) | 0.68 (0.44 - 1.05) | 1.58 (0.56 - 4.45) | 0.51 (0.09 - 2.76) |
| Southern Africa | 1.49 (1.12 - 2.00) | 1.54 (1.09 - 2.19) | - | - |
| **Level of education** |  |  |  |  |
| Master's degree and more | 1.00 | 1.00 | 1.00 | 1.00 |
| Bachelor's degree | 0.58 (0.43 - 0.77) | 0.57 (0.40 - 0.81) | 1.42 (0.64 - 3.17) | 0.79 (0.28 - 2.23) |
| Secondary/High School | 0.91 (0.62 - 1.36) | 0.82 (0.52 - 1.31) | 0.70 (0.14 - 3.40) | 0.81 (0.12 - 5.42) |
| Primary/Less | 0.22 (0.12 - 0.41) | 0.06 (0.01 - 0.25) | - | - |
| **Occupation** |  |  |  |  |
| Non-healthcare | 1.00 | 1.00 | 1.00 | 1.00 |
| Healthcare | 0.32 (0.25 - 0.40) | 0.30 (0.22 - 0.41) | 0.30 (0.14 - 0.67) | 0.59 (0.23 - 1.54) |
| **Working status** |  |  |  |  |
| Employed | 1.00 | 1.00 | 1.00 | 1.00 |
| Unemployed | 1.40 (1.06 - 1.84) | 1.06 (0.76 - 1.49) | 4.0 (0.55 - 36.83) | 7.60 (0.84 - 68.97) |
| **Marital/family status** |  |  |  |  |
| Married | 1.00 | 1.00 | 1.00 | 1.00 |
| Not married ‡ | 0.74 (0.58 - 0.95) | 0.76 (0.57 - 1.02) | 2.34 (1.02 - 5.41) | 2.54 (0.92 - 7.00) |
| **Religion** |  |  |  |  |
| Christians | 1.00 | 1.00 | 1.00 | 1.00 |
| Others | 1.10 (0.76 - 1.61) | 0.57 (0.34 - 0.97) | 0.79 (0.22 - 2.86) | 1.75 (0.43 - 7.18) |
| **Smoking status** |  |  |  |  |
| **Ex-smoker** | 1.00 | 1.00 | 1.00 | 1.00 |
| **Current smoker** | 1.21 (0.62 - 2.36) | 1.60 (0.71 - 3.58) | 1.54 (0.14 - 17.33) | 8.0 (0.31 - 206.37) |
| **Non-smoker** | 1.01 (0.62 - 1.65) | 1.12 (0.61 - 2.06) | 0.61 (0.18 - 2.00) | 3.18 (0.34 - 30.10) |
| **Have you been vaccinated for any condition** |  |  |  |  |
| **No** | 1.00 | 1.00 | 1.00 | 1.00 |
| **Yes** | 0.39 (0.26 - 0.57) | 0.46 (0.29 - 0.72) | 0.70 (0.18 - 2.76) | 0.75 (0.14 - 4.01) |
| **Any pre-existing conditions<sup>§</sup>** |  |  |  |  |
| **No** | 1.00 | 1.00 | 1.00 | 1.00 |
| **Yes** | 0.82 (0.60 - 1.12) | 0.84 (0.58 - 1.24) | 0.57 (0.26 - 1.25) | 0.44 (0.15 - 1.26) |
| **Knowledge** | 0.99 (0.97 - 1.01) | 1.03 (1.01 - 1.06) | 0.82 (0.73 - 0.91) | 0.87 (0.77 - 0.98) |
| **Attitude** | 0.96 (0.91 - 1.01) | 0.96 (0.90 - 1.03) | 0.99 (0.80 - 1.24) | 1.15 (0.90 - 1.46) |
| **Perception** | 0.88 (0.84 - 0.92) | 0.90 (0.85 - 0.94) | 0.77 (0.66 - 0.90) | 0.84 (0.70 - 1.01) |

If 95% confidence intervals (CI) around RRs that lies between 1.00 indicate not statistically significant. All comparisons were made against vaccinated pregnant women (RR=1.0).
‡ includes single, divorced, and widowed.
§includes the presence of any of the following conditions: cancer, diabetes, hypertension, asthma, kidney disease, any heart condition, sickle cell anemia

For those in diaspora, older age (>38years) [RR=0.13, 95% CI; 0.03 - 0.65], working in healthcare sector [RR=0.32, 95% CI; 0.25 – 0.40], having a more knowledge [0.82, 95% CI; 0.73 - 0.91] and better perception scores [RR=0.77, 95% CI; 0.66 – 0.90], were associated with lower risk of COVID-19 vaccine resistance, while not being married [RR=0.74, 95% CI; 0.58 – 0.95] had a higher risk of being resistant.

### Adjusted analysis of factors associated with COVID-19 vaccine uptake, resistance and hesitancy in SSA

Table 4 presents the associated factors of COVID-19 vaccine resistance and hesitancy in this study. After controlling for potential confounders in the local resident group, East African respondents were more likely to be resistant [ARR= 3.33, 95% CI: 1.40 - 7.94] and hesitant [ARR= 4.64, 95% CI; 1.84 - 11.70] towards receiving COVID-19 vaccines while Central African respondents were less likely to be resistant [ARR= 0.46, 95% CI; 0.32 - 0.68] or hesitant [ARR= 0.44, 95%CI: 0.27 - 0.72] towards the vaccines. Having a bachelor’s degree [ARR= 0.54, 95% CI; 0.38 - 0.76] or lower, being a health care worker [ARR= 0.24, 95% CI; 0.18 - 0.32], being previously vaccinated for any condition [ARR= 0.45, 95% CI; 0.30 - 0.69], and having a lower risk perception score [ARR= 0.86, 95% CI; 0.82 – 0.90] were associated with reduced risk of being resistant towards the COVID-19 vaccines among local residents in SSA. Among those in the diaspora, respondents who were aged 49 years and older [ARR= 0.17, 95% CI; 0.03 – 0.95], those who work in healthcare sectors [ARR=0.25, 95% CI; 0.10 - 0.62], as well as those with lower knowledge scores [ARR= 0.82, 95% CI; 0.73 – 0.91] were less likely to resist taking the COVID-19 vaccines.

**Table 4:**
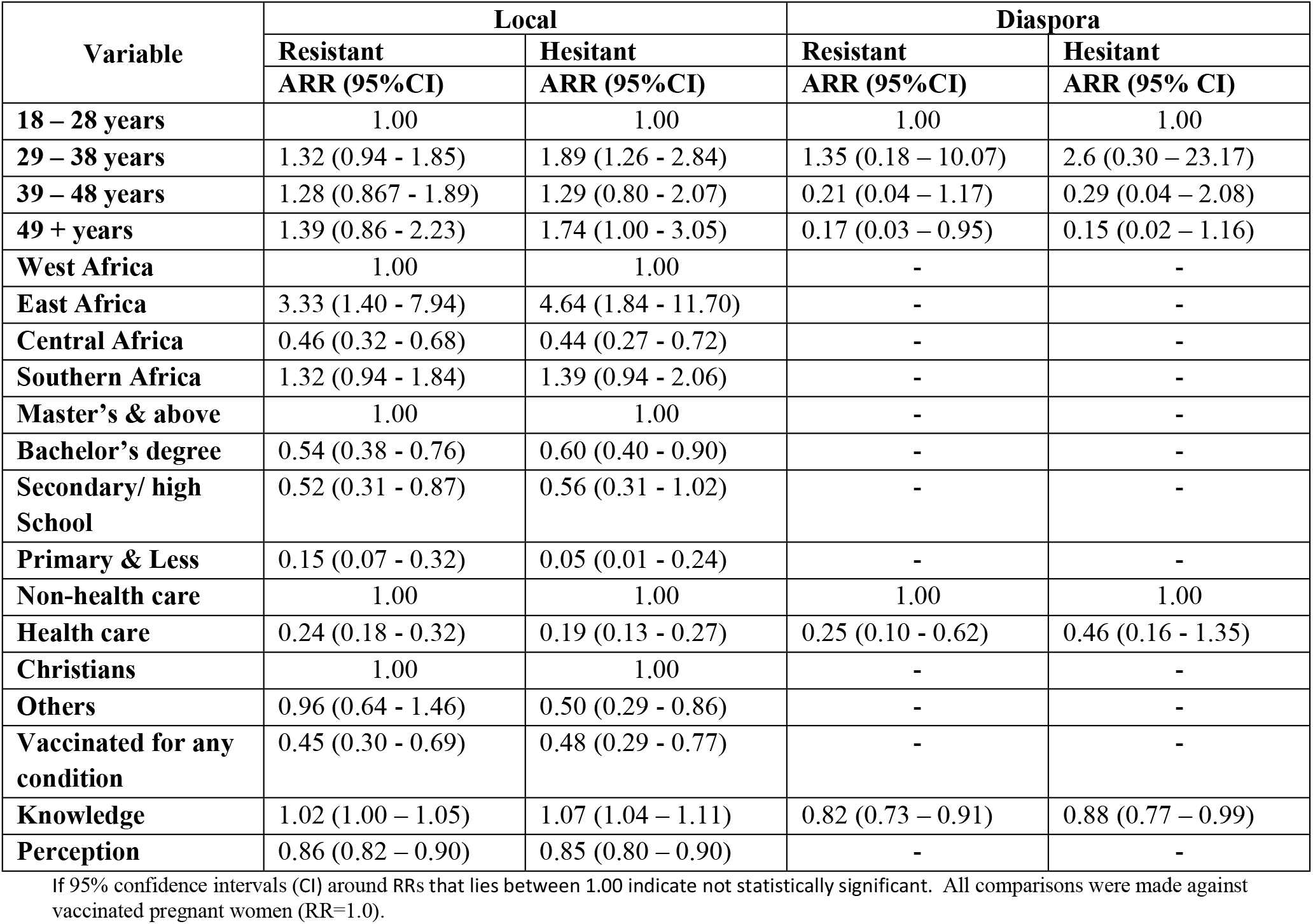
Adjusted Relative Risk (ARR) for factors associated with vaccine hesitancy among SSA residents living in (Locals) and outside of Africa (Diaspora). The base reference was COVID-19 vaccine uptake for all variables.

Regarding COVID-19 vaccine hesitancy among local residents in SSA, the significant factors included East and Central African origin, aged between 29 – 38 years, being a health care worker, having a bachelor’s degree or less, non-Christians, having been previously vaccinated for other conditions, higher knowledge and lower perception scores. While for those in diaspora being a health care worker [ARR= 0.46, 95% CI; 0.16 - 1.35] and having lower knowledge scores [ARR= 0.88, 95% CI; 0.77 – 0.99] were the factors that were significant for being hesitant.

## Discussion

The purpose of this study was to compare the uptake, resistance and hesitancy of the COVID-19 vaccine between the local residents and diaspora dwellers in SSA region of the African continent. Uptake of the COVID-19 vaccine was found to be twice as high among residents in the diaspora compared to local SSA residents. The WHO and Centers for Disease Control and Prevention (CDC) have suggested that the low vaccination rates in low-and-middle-income countries is in part, due to inequitable distribution of vaccines. Accessibility to vaccines may have played a role in the low uptake rates in our study. At the time of the study, half of the 52 African countries that had received vaccines had only vaccinated up to 2% of their population at the time of this study, and 15 countries had vaccinated up to 10%.[42] However, majority of those residing in Africa and the diaspora were either resistant or hesitant to get vaccinated. This finding is different from that reported in a previous study [43] where a higher proportion of African residents and those in the diaspora were willing to accept the vaccine when offered. A survey conducted by CDC Africa prior to the introduction of vaccines on the continent found that the willingness to take the vaccine in 15 African countries ranged from 59% to 93%,[44] which was in contrast with our findings of greater resistance towards COVID-19 vaccination. Studies conducted in the US and UK showed that Africans/Blacks were 13 times more likely to be hesitant than Whites [45-46] which is similar to the high proportions of SSA in diaspora who were either hesitant or resistant to taking COVID-19 vaccines.

Socio-demographic characteristics have been shown to play significant roles in vaccine hesitancy and resistance.[46] In this study, age, region of origin, educational level, occupation and religion were significantly associated with either vaccine hesitancy or resistance among local and diaspora residents. Younger age groups among the local residents were almost twice likely to be hesitant and older age groups were less likely to be resistant to vaccines. This finding is consistent with other previous studies,[32,37,46-47] and may also be related to the fact that COVID-19 is more likely to present in the severe form among older age groups, making them more likely to accept the vaccine for their protection.

Local East African respondents were three times more likely to resist and almost five times more likely to be hesitant than West Africans. This may be due to misinformation about COVID-19 [48] and its vaccines [49] which was reported to be more common in East African countries such as Tanzania. The results showed that the least educated respondents were less likely to be resistant or hesitant. This may be as a result of not comprehending the scientific arguments being advanced against the vaccines and having to make choices based on past experiences or the information they do understand. A recent study in the US showed a similar pattern with those with lower levels of education showing less hesitancy than those with higher.[49] This is contrary to the results obtained in other studies.[35-37,42-43] A statement by a 61 year old on Africa news may provide an insight into the mindset of those who are less educated thereby making them more likely to accept vaccination: “If in the time of our mothers, in the time we were little children if these “WhatsApp doctors” had existed (people who post unreliable medical information on social media) I think we would have all died because our mothers who did not go to school agreed to vaccinate us against smallpox, measles, polio -- all the other diseases without debate. Today, we are more educated, but curiously, we refuse vaccination. This is a certain danger for our society, according to what I have read here and there. The Congo is being blacklisted because we risk many deaths if we don’t accept vaccination”.[50]

Both local and diaspora healthcare workers showed less likelihood of being either resistant or hesitant as compared to non-healthcare workers in this study. Resistance and hesitancy have been found among health workers though lower when compared to non-healthcare workers.[50-55] However, Blacks /African health workers still show higher risk than their counterparts of being resistant/hesitant irrespective of the country they are in. Vaccine resistance and/or hesitancy is a hindrance to the vaccination campaign, as such, health workers who should be well educated about the vaccines are likely to exert an influence on others and possibly deter them from getting vaccinated. Most findings in the cited papers found that the fear of side effects was usually the reason for hesitancy and resistance among health workers. [51-53]

Among the local residents, individuals form other religions were less likely to be vaccine hesitant compared to those of the Christian faith. Religion has been reported to play a huge role in the life of Africans and influences their health seeking behavior.[57-58] Olagoke et al. reported that some religious views have contributed to the rejection of vaccination.[59] However, an intervention study conducted among American Christians,[60] showed that with proper presentation of scientific facts, such negative views can be changed. Community engagement with religious leaders has also been advocated as a means of addressing vaccine hesitancy.[61]

Local residents who had been previously vaccinated for other conditions were less likely to be COVID-19 vaccine resistant or hesitant. This finding emphasizes the influence of past experiences which can build confidence in the efficacy of vaccines. Other studies have also shown a willingness to be vaccinated among those who had previously received vaccinations for other diseases such as flu, yellow fever, hepatitis.[62-63] Knowledge of COVID-19 vaccine was a significant factor among both local and diaspora residents. Knowledge has been shown to reduce resistance to vaccine acceptance. Africans in the diaspora were less likely to be hesitant or resistant to vaccines as compared to their counterparts residing in Africa. This may still be related to misinformation and the need for health messages to be relayed in the languages familiar to the people. Recent studies have shown a decline in those who are hesitant and this has been attributed to the availability of accurate information that reduces fear and leads to making informed decisions.[64] Exposure to accurate information and increased knowledge about COVID-19 vaccines may help those who are hesitant to be more receptive to vaccines. Among local residents, higher perception scores showed a lower odd of being either resistant or hesitant. The perception that one is likely to be at risk of contracting a disease can result in people taking appropriate measures to protect themselves from contracting the disease.

### Strengths and limitations

This is the first large scale study to compare acceptance of COVID-19 vaccines between sub-Saharan African local residents and those in the diaspora. The study employed robust analyses to control for potential confounders to reduce the possibility of a bias. The distribution of the questionnaire in both English and French languages using an internet-based methodology, which was the only reliable means to disseminate information at the time of this study to a wider audience. Notwithstanding these strengths, the study has some limitations. For example, the study did not explore concerns about vaccine safety which may be an important determinant of vaccine hesitancy. The cross-sectional nature of the study means that causation cannot be determined. The survey was distributed electronically using social media platforms and emails, and this may have inadvertently excluded some potential participants whose opinions may have differed, such as those without internet access and people living in rural areas, where internet penetration remains relatively low.[65] The survey was presented in English and French and thus inadvertently excluding some of the Portuguese or Arabic-speaking SSA countries from participating. Although the study showed satisfactory internal validity, its generalization or transferability to all SSA countries may be limited.

## Conclusion

The study showed that Africans residing both locally and in diaspora are mostly either resistant or hesitant to the COVID-19 vaccines. Factors that influenced resistance and hesitancy among local residents included younger age, being from East and Central Africa, lower levels of education, history of previous vaccinations, being a health care worker, knowledge and perceptions of COVID-19 vaccine. For Africans in the diaspora, being hesitant or resistant to COVID-19 vaccines are influenced by older age, being a health care worker and having adequate knowledge of vaccines. Appropriate interventions such as public health messaging are required to enhance COVID-19 uptake to achieve sufficient vaccine coverage.

## Data Availability

Data for this study has been included in the submission

## Acknowledgement

None to acknowledge.

## Supporting information

S1 Table. Sample of the survey items used in this study

## References

1. Liu YC, Kuo RL, Shih SR. COVID-19: The first documented coronavirus pandemic in history. Biomed J. 2020 Aug; 43(4):328–33. doi: 10.1016/j.bj.2020.04.007. Epub 2020 May 5. PubMed PMID: 32387617; PubMed Central PMCID: PMC7199674.

2. Seong H, Hyun HJ, Yun JG, Noh JY, Cheong HJ, Kim WJ et al. Comparison of the second and third waves of the COVID-19 pandemic in South Korea: Importance of early public health intervention. Int J Infect Dis. 2021 Mar; 104:742–5. doi: 10.1016/j.ijid.2021.02.004. Epub 2021 Feb 5. PubMed PMID: 33556610; PubMed Central PMCID: PMC7863747.

3. Salyer SJ, Maeda J, Sembuche S, Kebede Y, Tshangela A, Moussif M et al. The first and second waves of the COVID-19 pandemic in Africa: a cross-sectional study. Lancet. 2021 Apr 3; 397(10281):1265–75. doi: 10.1016/S0140-6736(21)00632-2. Epub 2021 Mar 24. PubMed PMID: 33773118; PubMed Central PMCID: PMC8046510.

4. Maslo C, Friedland R, Toubkin M, Laubscher A, Akaloo T, Kama B. Characteristics and outcomes of hospitalized patients in South Africa during the COVID-19 Omicron wave compared with previous waves. JAMA. 2022 Feb 8; 327(6):583–4. doi: 10.1001/jama.2021.24868. PubMed PMID: 34967859; PubMed Central PMCID: PMC8719272.

5. Grabowski F, Kochańczyk M, Lipniacki T. The spread of SARS-CoV-2 variant Omicron with a doubling time of 2.0–3.3 days can be explained by immune evasion. Viruses. 2022; 14(2):294. https://doi.org/10.3390/v14020294.

6. Brandal LT., MacDonald E, Veneti L, Ravlo T, Lange H, Naseer U et al. Outbreak caused by the SARS-CoV-2 Omicron variant in Norway, November to December 2021. Euro Surveill. 2021; 26 (50). pii=2101147. https://doi.org/10.2807/1560-7917.ES.2021.26.50.2101147.

7. Helmsdal G, Hansen OK, Møller LF, Christiansen DH, Petersen MS, Kristiansen MF. Omicron outbreak at a private gathering in the Faroe Islands, infecting 21 of 33 triple-vaccinated healthcare workers. Clin Infect Dis. 2022 Feb 3:ciac089. doi: 10.1093/cid/ciac089. Epub ahead of print. PubMed PMID: 35134167.

8. Saxena SK, Kumar S, Ansari S, Paweska JT, Maurya VK, Tripathi AK et al. Characterization of the novel SARS-CoV-2 Omicron (B.1.1.529) variant of concern and its global perspective. J Med Virol. 2022 Apr; 94(4):1738–44. doi: 10.1002/jmv.27524. Epub 2022 Jan 11. Pub Med PMID: 34905235.

9. Odusanya OO, Odugbemi BA, Odugbemi TO, Ajisegiri WS. COVID-19: A review of the effectiveness of non-pharmacological interventions. Niger Postgrad Med J. 2020; 27(4)261-7.

10. Gokmen Y, Baskici C, Ercil Y. Effects of non-pharmaceutical interventions against COVID-19: A cross-country analysis. Int J Health Plann Manage. 2021 Jul; 36(4):1178–88. doi: 10.1002/hpm.3164. Epub 2021 Apr 5. PubMed PMID: 33819370; PubMed Central PMCID: PMC8251211.

11. Schuchat A. Human vaccines and their importance to public health. Procedia Vaccinol 2011; 5: 120–6.

12. World Health Organization [Internet]. Draft landscape and tracker of COVID-19 candidate vaccines; c2020 [cited 2022 Jan 23]. Available from: https://www.who.int/docs/default-source/a-future-for-children/novel-coronavirus_landscape_covid-19.pdf?sfvrsn=4d8bd201_1.

13. World Health Organization [Internet]. Guidance document: Status of COVID-19 Vaccines within WHO EUL/PQ evaluation process; c2022 [cited 2021 May 20]. Available from: https://extranet.who.int/pqweb/sites/default/files/documents/Status_COVID_VAX_02March2022.pdf

14. National Research Council (US) Division of Health Promotion and Disease Prevention. Vaccine Supply and Innovation. Washington (DC): National Academies Press (US); c1985 [cited 2021 may 21] 3, Vaccine Availability: Concerns, Barriers, and Impediments. Available from: https://www.ncbi.nlm.nih.gov/books/NBK216807/.

15. Valckx S, Crèvecoeur J, Verelst F, Vranckx M, Hendrickx G, Hens N et al. Individual factors influencing COVID-19 vaccine acceptance in between and during pandemic waves (July-December 2020). Vaccine. 2022 Jan 3; 40(1):151–61. doi: 10.1016/j.vaccine.2021.10.073. Epub 2021 Dec 1. Pubmed PMID: 34863621; PubMed Central PMCID: PMC8634074..

16. Mustapha M, Lawal BK, Sha’aban A, Jatau AI, Wada AS, Bala AA et al. Factors associated with acceptance of COVID-19 vaccine among University health sciences students in Northwest Nigeria. PLoS One. 2021 Nov 29;16(11):e0260672. doi: 10.1371/journal.pone.0260672. PubMed PMID: 34843594; PubMed Central PMCID: PMC8629299.

17. Kamal AHM, Sarkar T, Khan MM, Roy SK, Khan SH, Hasan SMM et al. Factors affecting willingness to receive COVID-19 vaccine among adults: a cross-sectional study in Bangladesh. Journal of Health Management.2021 Oct. doi: 10.1177/09735984211050691.

18. Olliaro P, Torreele E, Vaillant M. COVID-19 vaccine efficacy and effectiveness-the elephant (not) in the room. Lancet Microbe. 2021 Jul; 2(7):e279–e280. doi: 10.1016/S2666-5247(21)00069-0. Epub 2021 Apr 20. PubMed PMID: 33899038; PubMed Central PMCID: PMC8057721.

19. Cho W, Kirwin MF [Internet]. WP71: A vicious circle of corruption and mistrust in institutions in sub-Saharan Africa: a micro-level analysis. Afrobarometer; c2007 [cited 2022 Jan 23]. Available from: https://afrobarometer.org/publications/wp71-vicious-circle-corruption-and-mistrust-institutions-sub-saharan-africa-micro-level.

20. Razai MS, Chaudhry U A R, Doerholt K, Bauld L, Majeed A. Covid-19 vaccination hesitancy BMJ. 2021; 373:n1138. doi:10.1136/BMJ.n1138.

21. World Health Organization [Internet]. Ten threats to global health in 2019; c2019 [cited 2022 Jan 23]. Available from: https://www.who.int/news-room/spotlight/ten-threats-to-global-health-in-2019.

22. Geoghegan S, O’Callaghan KP, Offit PA. Vaccine safety: myths and misinformation. Front Microbiol. 2020 Mar 17; 11:372. doi: 10.3389/fmicb.2020.00372. PubMed PMID: 32256465; PubMed Central PMCID: PMC7090020.

23. Umakanthan S, Patil S, Subramaniam N, Sharma R. COVID-19 vaccine hesitancy and resistance in India explored through a population-based longitudinal survey. Vaccines (Basel). 2021 Sep 24; 9(10):1064. doi: 10.3390/vaccines9101064. PubMed PMID: 34696172; PubMed Central PMCID: PMC8537475.

24. Khubchandani J, Sharma S, Price JH, Wiblishauser MJ, Sharma M, Webb FJ. COVID-19 vaccination hesitancy in the United States: a rapid national assessment. J Community Health. 2021 Apr; 46(2):270–7. doi: 10.1007/s10900-020-00958-x. Epub 2021 Jan 3. PubMed PMID: 33389421; PubMed Central PMCID: PMC7778842.

25. Kini A, Morgan R, Kuo H, Shea P, Shapiro J, Leng SX et al. Differences and disparities in seasonal influenza vaccine, acceptance, adverse reactions, and coverage by age, sex, gender, and race. Vaccine. 2022 Mar 8; 40(11):1643–54. doi: 10.1016/j.vaccine.2021.04.013. Epub 2021 Apr 28. PubMed PMID: 33933316; PubMed Central PMCID: PMC8551304.

26. Niederhauser VP, Stark M. Narrowing the gap in childhood immunization disparities. Pediatr Nurs. 2005 Sep-Oct; 31(5):380-6. PubMed PMID: 16295153.

27. Webb Hooper M, Nápoles AM, Pérez-Stable EJ. No populations left behind: vaccine hesitancy and equitable diffusion of effective COVID-19 vaccines. J Gen Intern Med. 2021 Jul; 36(7):2130–3. doi: 10.1007/s11606-021-06698-5. Epub 2021 Mar 22. PubMed PMID: 33754319; PubMed Central PMCID: PMC7985226.

28. Malik AA, McFadden SM, Elharake J, Omer SB. Determinants of COVID-19 vaccine acceptance in the US. EClinicalMedicine. 2020 Sep; 26:100495. doi: 10.1016/j.eclinm.2020.100495. Epub 2020 Aug 12. PubMed PMID: 32838242; PubMed Central PMCID: PMC7423333.

29. The ICD “Experience Africa” Program [Internet]. The African Diaspora. Introduction to the African Diaspora across the World; c2021 [cited 2022 Jan 23]. Available from: https://www.experience-africa.de/index.php?en_the-african-diaspora.

30. African Diaspora [Internet]. Meeting of Experts on the Definition of the African Diaspora 11 – 12 April 2005 Addis Ababa, Ethiopia. Report of the meeting of experts from members of States on the definition of African Diaspora; c2021 [cited 2022 Jan 23]. Available from: http://www.dirco.gov.za/diaspora/definition.html.

31. Riesen M, Konstantinoudis G, Lang P, Low N, Hatz C, Maeusezahl M et al. Exploring variation in human papillomavirus vaccination uptake in Switzerland: a multilevel spatial analysis of a national vaccination coverage survey. BMJ Open. 2018 May 17; 8(5):e021006. doi: 10.1136/bmjopen-2017-021006. PubMed PMID: 29773702; PubMed Central PMCID: PMC5961588.

32. McAuslane H, Utsi L, Wensley A, Coole L, Inequalities in maternal pertussis vaccination uptake: a cross-sectional survey of maternity units, J Public Health. 2018 Mar; 40(1):121–8. doi: https://doi.org/10.1093/pubmed/fdx032.

33. Basak P, Abir T, Al Mamun A, Zainol NR, Khanam M, Haque MR et al. A global study on the correlates of Gross Domestic Product (GDP) and COVID-19 vaccine distribution. Vaccines. 2022; 10(2):266. doi: https://doi.org/10.3390/vaccines10020266.

34. World Health Organization [Internet]. Africa on track to control COVID-19 pandemic in 2022; c2022 [cited 2022 Jan 23]. Available from: https://www.afro.who.int/news/africa-track-control-covid-19-pandemic-2022.

35. World Health Organization Regional Office for Africa [Internet]. Risks and challenges in Africa’s COVID-19 vaccine rollout; c2021 [cited 2022 Jan 23] Available from: https://www.afro.who.int/news/risks-and-challenges-africas-covid-19-vaccine-rollout.

36. Dzinamarira T, Nachipo B, Phiri B, Musuka G. COVID-19 vaccine roll-out in South Africa and Zimbabwe: urgent need to address community preparedness, fears and hesitancy. Vaccines. 2021; 9(3):250. doi: https://doi.org/10.3390/vaccines9030250.

37. Abu EK, Oloruntoba R, Osuagwu UL, Bhattarai D, Miner CA, Goson PC et al. Risk perception of COVID-19 among sub-Sahara Africans: a web-based comparative survey of local and diaspora residents. BMC Public Health. 2021 Aug 18; 21(1):1562. doi: 10.1186/s12889-021-11600-3. PubMed PMID: 34404377; PubMed Central PMCID: PMC8370831.

38. World Medical Assembly [Internet]. WMA Declaration of Helsinki - Ethical Principles for Medical Research Involving Human Subjects. c2013 [cited 2022 Jan 23]. Available from: https://www.wma.net/policies-post/wma-declaration-of-helsinki-ethical-principles-for-medical-research-involving-human-subjects/.

39. Geldsetzer P. Use of rapid online surveys to assess people’s perceptions during infectious disease outbreaks: a cross-sectional survey on COVID-19. J Med Internet Res. 2020 Apr 2; 22(4):e18790. doi: 10.2196/18790. Pubmed PMID: 32240094; PubMed Central PMCID: PMC7124956.

40. Biasio LR, Bonaccorsi G, Lorini C, Pecorelli S. Assessing COVID-19 vaccine literacy: a preliminary online survey. Hum Vaccin Immunother. 2021 May 4;17(5):1304–12. doi: 10.1080/21645515.2020.1829315. Epub 2020 Oct 29. PubMed PMID: 33118868; PubMed Central PMCID: PMC8078752.

41. Buzasi K. Linguistic Situation in Twenty sub-Saharan African Countries: A Survey-based Approach. Afr Stud. 2016; 75:358–80. doi: 10.1080/00020184.2016.1193376

42. World Health Organization Regional Office for Africa [Internet]. Fifteen African countries hit 10% COVID-19 vaccination goal; c2021 [cited 2022 Jan 23]. Available from: https://www.afro.who.int/news/fifteen-african-countries-hit-10-covid-19-vaccination-goal.

43. Anjorin AA, Odetokun IA, Abioye AI, Elnadi H, Umoren MV, Damaris BF, et al. Will Africans take COVID-19 vaccination? PLoS ONE. 2021; 16(12): e0260575. doi: https://doi.org/10.1371/journal.pone.0260575.

44. Africa Centers for Disease Control and Prevention [Internet]. COVID 19 vaccine perceptions: A 15 country study; c2021 [cited 2022 Jan 23]. Available from: https://africacdc.org/download/covid-19-vaccine-perceptions-a-15-country-study/.

45. Momplaisir FM, Kuter BJ, Ghadimi F, Browne S, Nkwihoreze H, Feemster KA et al. Racial/ethnic differences in COVID-19 vaccine hesitancy among health care workers in 2 large academic hospitals. JAMA Netw Open. 2021 Aug 2; 4(8):e2121931. doi: 10.1001/jamanetworkopen.2021.21931. PubMed PMID: 34459907; PubMed Central PMCID: PMC8406078.

46. Robertson E, Reeve KS, Niedzwiedz CL, Moore J, Blake M, Green M et al. Predictors of COVID-19 vaccine hesitancy in the UK household longitudinal study. Brain Behav Immun. 2021 May; 94:41–50. doi: 10.1016/j.bbi.2021.03.008. Epub 2021 Mar 11. PubMed PMID: 33713824; PubMed Central PMCID: PMC7946541.

47. Lazarus JV, Wyka K, Rauh L, Rabin K, Ratzan S, Gostin LO et al. Hesitant or not? The association of age, gender, and education with potential acceptance of a COVID-19 vaccine: a country-level analysis. J Health Commun. 2020 Oct 2;25(10):799–807. doi: 10.1080/10810730.2020.1868630. PubMed PMID: 33719881.

48. Osuagwu UL, Miner CA, Bhattarai D, Mashige KP, Oloruntoba R, Abu EK et al. Misinformation about COVID-19 in sub-Saharan Africa: evidence from a cross-sectional survey. Health Secure. 2021 Jan-Feb;19(1):44-56. DOI: 10.1089/HS.2020.0202. PMID: 33606572.

49. King WC, Rubinstein M, Reinhart A, Mejia R. Time trends, factors associated with, and reasons for COVID-19 vaccine hesitancy: A massive online survey of US adults from January-May 2021. PLoS One. 2021 Dec 21; 16(12):e0260731. doi: 10.1371/journal.pone.0260731. PubMed PMID: 34932583; PubMed Central PMCID: PMC8691631.

50. Asala K; Africanews [Internet]. DR Congo: COVID-19 vaccine hesitation amid surging cases in third wave. [cited 2022 Feb 21]. Available from: https://www.africanews.com/2021/07/02/dr-congo-covid-19-vaccine-hesitation-amid-surging-cases-in-third-wave//.

51. Toth-Manikowski SM, Swirsky ES, Gandhi R, Piscitello G. COVID-19 vaccination hesitancy among health care workers, communication, and policy-making. Am J Infect Control. 2022 Jan; 50(1):20–5. doi: 10.1016/j.ajic.2021.10.004. Epub 2021 Oct 13. PubMed PMID: 34653527; PubMed Central PMCID: PMC8511871.

52. Amuzie CI, Odini F, Kalu KU, Izuka M, Nwamoh U, Emma-Ukaegbu U et al. COVID-19 vaccine hesitancy among healthcare workers and its socio-demographic determinants in Abia State, Southeastern Nigeria: a cross-sectional study. Pan Afr Med J. 2021 Sep 3; 40:10. doi: 10.11604/pamj.2021.40.10.29816. PubMed PMID: 34650660; PubMed Central PMCID: PMC8490164..

53. Aoun AH, Aon MH, Alshammari AZ, Moussa SA. COVID-19 vaccine hesitancy among health care workers in the Middle East Region. Open Public Health J. 2021; 14: 352–9. doi: 10.2174/1874944502114010352.

54. Angelo AT, Alemayehu DS, Dachew AM. Health care workers intention to accept COVID-19 vaccine and associated factors in southwestern Ethiopia. 2021. PLoS One. 2021 Sep 3; 16(9):e0257109. doi: 10.1371/journal.pone.0257109. PubMed PMID: 34478470; PubMed Central PMCID: PMC8415602.

55. Ashok N, Krishnamurthy K, Singh K, Rahman S, Majumder MAA. High COVID-19 Vaccine Hesitancy Among Healthcare Workers: Should Such a Trend Require Closer Attention by Policymakers? Cureus. 2021 Sep 15; 13(9):e17990. doi: 10.7759/cureus.17990. PubMed PMID: 34667668; PubMed Central PMCID: PMC8519358.

56. Algabbani F, Algabbani A. Vaccine hesitancy among healthcare providers during COVID-19 pandemic. Eur J Public Health. 2021; 31 Suppl_7. doi: https://doi.org/10.1093/eurpub/ckab164.101

57. Wirsiy FS, Nkfusai CN, Ako-Arrey DE, Dongmo EK, Manjong FT, Cumber SN. Acceptability of COVID-19 Vaccine in Africa. Int J MCH AIDS. 2021; 10(1):134–8. doi: 10.21106/ijma.482. Epub 2021 Apr 8. PubMed PMID: 33868778; PubMed Central PMCID: PMC8039868.

58. Costa JC, Weber AM, Darmstadt GL, Abdalla S, Victora CG. Religious affiliation and immunization coverage in 15 countries in Sub-Saharan Africa. Vaccine. 2020 Jan 29; 38(5):1160–9. doi: 10.1016/j.vaccine.2019.11.024. Epub 2019 Nov 30. PubMed PMID: 31791811; PubMed Central PMCID: PMC6995994.

59. Olagoke AA, Olagoke OO, Hughes AM. Intention to vaccinate against the novel 2019 coronavirus disease: the role of health locus of control and religiosity. J Relig Health. 2021 Feb; 60(1):65–80. doi: 10.1007/s10943-020-01090-9. Epub 2020 Oct 30. PubMed PMID: 33125543; PubMed Central PMCID: PMC7596314.

60. Chu J, Pink SL, Willer R. Religious identity cues increase vaccination intentions and trust in medical experts among American Christians. Proc Natl Acad Sci U S A. 2021 Dec 7; 118(49):e2106481118. doi: 10.1073/pnas.2106481118. PubMed PMID: 34795017; PubMed Central PMCID: PMC8670469.

61. Melillo S, Fountain D, Bormet M, O’Brien CJ; MOMENTUM Country and Global Leadership. Effects of faith actor engagement in the uptake and coverage of immunization in low- and middle-income countries (LMICs)* Phase 1 Global Landscape : Evidence Summary. c2021 [cited 2022 Jan 24]. Available from: https://usaidmomentum.org/wp-content/uploads/2021/07/Faith-Engagement-in-Immunization_Global-Landscape-Analysis_Evidence-Summary-Report_June-2021_Sec508comp-low.pdf.

62. El-Elimat T, AbuAlSamen MM, Almomani BA, Al-Sawalha NA, Alali FQ. Acceptance and attitudes toward COVID-19 vaccines: A cross-sectional study from Jordan. PLoS One. 2021 Apr 23; 16(4):e0250555. doi: 10.1371/journal.pone.0250555. PubMed PMID: 33891660; PubMed Central PMCID: PMC8064595.

63. Wake AD. The willingness to receive COVID-19 vaccine and its associated factors: “Vaccination refusal could prolong the war of this pandemic” -a systematic review. Risk Manag Healthc Policy. 2021 Jun 21; 14:2609–2623. doi: 10.2147/RMHP.S311074. PubMed PMID: 34188572; Pubmed Central PMCID: PMC8232962.

64. Siegler AJ, Luisi N, Hall EW, Bradley H, Sanchez Z, Lopman BA et al. Trajectory of COVID-19 vaccine hesitancy over time and association of initial vaccine hesitancy with subsequent vaccination. JAMA Netw Open. 2021; 4(9): e2126882–e2126882. doi:10.1001/jamanetworkopen.2021.26882.

65. Hjort, J, Poulsen, J. The arrival of fast internet and employment in Africa. American Economic Review. 2019; 109(3):1032–79. doi: 10.1257/aer.20161385

